# Association between corticosteroids and intubation or death among patients with COVID-19 pneumonia in non-ICU settings: an observational study using of real-world data from 51 hospitals in France and Luxembourg

**DOI:** 10.1101/2020.09.16.20195750

**Authors:** The COCORICO Collaborative Group, Viet-Thi Tran, Matthieu Mahévas, Firouze Bani-Sadr, Olivier Robineau, Thomas Perpoint, Elodie Perrodeau, Laure Gallay, Philippe Ravaud, François Goehringer, François-Xavier Lescure

**Affiliations:** Centre d’Epidémiologie Clinique, Hôpital Hôtel-Dieu, Assistance Publique-Hôpitaux de Paris, 75004 Paris, France; Université de Paris, CRESS, INSERM, INRA, F-75004 Paris, France; Service de Médecine Interne, Hôpital Henri-Mondor, Assistance Publique-Hôpitaux de Paris, 75000 Paris France; Université Paris-Est Créteil, 94000 Créteil, France; Service de Médecine Interne, Centre Hospitalier Universitaire de Reims, 51100 Reims, France; Service Universitaire des Maladies Infectieuses et du Voyageur, centre hospitalier, hôpital Guy Chatiliez, 59200 Tourcoing, France; Service des maladies infectieuses et tropicales, Hospices Civils de Lyon, 69004 Lyon, France; Service de Médecine Interne, Hospices Civils de Lyon, 69004 Lyon, France; Service de Maladies Infectieuses et Tropicales, Centre Hospitalier Régional Universitaire de Nancy, 54511 Vandoeuvre lès Nancy, France; Service de maladies infectieuses et tropicales, Hôpital Bichat, Assistance Publique Hôpitaux de Paris, 75018 Paris, France

**Keywords:** COVID-19, Corticosteroids, therapeutic evaluation, observational study

## Abstract

**Objective:** To assess the effectiveness of corticosteroids on outcomes of patients with mild COVID-19 pneumonia.

**Methods:** We used routine care data from 51 hospitals in France and Luxembourg to assess the effectiveness of corticosteroids at 0.8 mg/kg/day eq. prednisone (CTC group) vs standard of care (no-CTC group) among patients ≤ 80 years old with COVID-19 pneumonia requiring oxygen without mechanical ventilation. The primary outcome was intubation or death at Day 28. Baseline characteristics of patients were balanced using propensity score inverse probability of treatment weighting.

**Results:** Among the 891 patients included in the analysis, 203 were assigned to the CTC group. At day 28, corticosteroids did not reduce the rate of the primary outcome (wHR 0.92, 95% CI 0.61 to 1.39) nor the cumulative death rate (wHR 1.03, 95% CI 0.54 to 1.98). Corticosteroids significantly reduced the rate of the primary outcome for patients requiring oxygen ≥ at 3L/min (wHR 0.50, 95% CI 0.30 to 0.85) or C-Reactive Protein (CRP) ≥ 100mg/L (wHR 0.44, 95%CI 0.23 to 0.85). We found a higher number of hyperglycaemia events among patients who received corticosteroids, but number of infections were similar across the two groups.

**Conclusions:** We found no association between the use of corticosteroids and intubation or death in the broad population of patients ≤80 years old with COVID-19 hospitalized in non-ICU settings. However, the treatment was beneficial for patients with ≥ 3L/min oxygen or CRP ≥ 100mg/L at baseline. These data support the need to confirm the right timing of corticosteroids for patients with mild COVID.

**Short summary:** We assessed the effectiveness of corticosteroids among patients ≤ 80 years old with COVID-19, in non-ICU settings. Our results support the use of corticosteroids for patients receiving oxygen at ≥3L/min or with a C-reactive protein ≥ 100mg/L at baseline.

## Introduction

The coronavirus disease 2019 (COVID-19) is a life-threatening disease that can cause fatal pneumonia. COVID-19 pneumonia is associated with a hyper-inflammation phase, which is deemed responsible for the clinical worsening of many patients [1,2].

At the onset of the COVID-19 pandemic, guidance regarding corticosteroids for patients without acute respiratory distress syndrome (ARDS) was mixed. For example, in April 2020, guidelines from the Infectious Diseases Society of America issued a weak recommendation against corticosteroids, except for patients with COVID-19 and ARDS treated in the context of a clinical trial [3]. During the main phase of the pandemic in Europe (March to June 2020), only evidence from small observational studies, with contrasting results, was available [4–11]. In July 2020, results of the RECOVERY trial were published, showing that the use of dexamethasone reduced the 28-day mortality, in patients receiving oxygen without invasive mechanical ventilation and in those receiving mechanical ventilation at the time of randomization [12]. Moreover, two other recent trials have suggested the efficacy of hydrocortisone and dexamethasone on the outcomes of critically ill patients [13–15]. A meta-analysis of seven trials that evaluated the efficacy of corticosteroids in critically ill patients with COVID-19 who received dexamethasone or hydrocortisone showed a reduction in 28-days mortality [16]. Yet, several questions remain unanswered. First, for patients with mild COVID hospitalized in non-ICU settings and receiving oxygen without mechanical ventilation, only the results from the RECOVERY trial are available (which showed an overall benefit for patients who were receiving oxygen but not for those without oxygen at the time of randomization). Unfortunately, these preliminary results do not provide sufficient information on a potential heterogeneity of the treatment effect in this broad population. Second, the harms of using of corticosteroids for COVID-19 in this population have not been thoroughly explored.

## Methods

### Study design

We used data collected from routine care to assess the efficacy and safety of corticosteroid therapy (at least 0.8 mg prednisone-equivalent per kilogram body weight) for patients hospitalised with a COVID-19 infection, requiring oxygen and with an inflammatory syndrome.

### Setting

Our study involved the internal medicine or infectious disease wards from 51 hospitals in France and Luxembourg. Among them, 25 were from university hospitals and 26 were from general hospitals. All centres were part of a network coordinated by REACTing (INSERM) against COVID-19 set-up after the first SARS-CoV-2 infected patients diagnosed in France on January 25^th^, 2020.

### Study population

Physicians performed a patient-by-patient screening of all patients hospitalised between March, 1^st^ and May,1^st^, 2020 and included all consecutive patients aged between 18 and 80 years, who had a PCR-confirmed SARS-CoV-2 infection, an inflammatory syndrome with a C-reactive protein (CRP) level ≥ 40mg/L, and required oxygen by mask or nasal prongs (corresponding to a WHO progression score of 5).

Exclusion criteria were 1) the presence of a contraindication to corticosteroids; 2) the start of corticosteroids before hospitalisation; 3) deep undernutrition with a body mass index (BMI) <16; 4) renal diseases requiring dialysis; 5) chronic heart failure NYHA IV; 6) liver cirrhosis Child C, 7) chronic respiratory insufficiency under oxygen therapy before admission; 8) start of a treatment with anti-interleukin drugs (e.g. tocilizumab) before or concomitantly to the start of corticosteroids; 9) organ failure requiring immediate admission to the intensive care unit (ICU) or continuous care unit (CCU) (incl. patients in requiring non-invasive ventilation with provision of positive airway pressure [15]); 11) discharge from the ICU to standard care;12) decision to limit and stop active treatments; and 13) inclusion in the DISCOVERY trial (NCT04315948).

The study was performed in accordance with the declaration of Helsinki. It received approval by the IRB of the Henri-Mondor Hospital (AP-HP), France (number: 00011558) and by the National ethics committee of Luxembourg (number: 0620-101). The study was based on data from routine care already collected at the time of the study; thus informed consent of participants was not required. All patients were informed that their hospital data would be used for research purposes and could refuse such use of their data.

### Treatment strategies

We compared two treatment strategies, the initiation of corticosteroids with at least 0.8 mg/kg/day eq. prednisone or 0,4 mg/kg/day eq. prednisone if co-administrated with lopinavir-ritonavir (CTC group) versus the standard of care (no CTC group). These values were chosen to account for dose rounding by physicians who had the intent to treat patients with corticosteroids at 1 mg/kg/day and 0.5 mg/kg/day eq. prednisone. Lower dose of corticosteroids when associated with Lopinavir-ritonavir aimed at accounting a potential drug-drug interaction between ritonavir and steroids[17]. Standard of care consisted of supportive therapy and on treating the symptoms to prevent respiratory failure. No systematic antibiotic prophylaxis nor antivirals were provided to patients in the study centres.

To emulate a pragmatic trial, patients in the CTC group could start corticosteroids within a “grace period” of 5 days after eligibility. The grace period would correspond, in the target trial to the time during which patients assigned to a given treatment strategy and who initiated the treatment slightly after time zero are still considered compliant with the protocol. The grace period ensures that the strategies remain realistic but also increases the number of people in the observational database whose data can be used to emulate the target trial. The definition of group assignment based on patients’ observational data is reported in **Supplementary material 1**.

### Start and end of follow-up

The start of follow-up (baseline or time zero) for each individual was the time all eligibility criteria (oxygen therapy and inflammatory syndrome with a CRP level ≥ 40mg/L) were checked. All patients were followed up from baseline until whichever of the following events occurred first: (1) death, (2) loss to follow-up, or (3) end of follow-up, which occurred at least 28 days after baseline.

### Outcomes

The primary outcome was intubation or death at Day 28. Secondary outcomes — all at Day 28 — were death from any cause, weaning from oxygen, and discharge from hospital to home/rehabilitation. Patient discharged who were secondarily readmitted in hospital and who were hospitalised at Day 28, whatever the reason, were not considered discharged from hospital at that day. All adverse events were abstracted from electronic health records in free text and independently recoded by four physicians (XL, FG, TP and MM). In case of disagreement, consensus was obtained.

### Statistical analysis

Our causal contrast of interest was the per-protocol effect. We compared participants who received corticosteroids with at least 0.8 mg/kg/day eq. prednisone or 0,4 mg/kg/day eq. prednisone if co-administrated with lopinavir-ritonavir (CTC group) within 5 days from eligibility to those who did not receive the drug.

Our primary analysis aimed at evaluating the average treatment effect (ATE) of corticosteroids in the whole population [18]. We used an inverse probability of treatment weighting (IPTW) approach based on patients’ propensity score (i.e., patients’ predicted probability of receiving a certain CTC given their baseline covariates) to balance the differences in baseline variables between treatment groups.^11,12^ A non-parsimonious multivariable logistic regression model was constructed to estimate each patient’s propensity score. Variables of the propensity score (PS) model were planned and prespecified before any outcome analyses, and included age; gender; presence of chronic respiratory insufficiency or a chronic respiratory pathology likely to decompensate during a viral infection; heart failure [NYHA I, II or III]; chronic kidney disease; liver cirrhosis (with Child-Pugh class A or B); personal history of cardiovascular disease [hypertension, stroke, coronary artery disease, or cardiac surgery]; insulin-dependent diabetes mellitus, or diabetic microangiopathy or macroangiopathy; immunosuppression (because of immunosuppressive drugs, including anticancer chemotherapy; uncontrolled HIV infection or HIV infection with CD4 cell counts < 200/µl; or a haematological malignancy); BMI (≥30 kg/m^2^ or not); treatment by angiotensin-converting enzyme inhibitors (ACEIs) or angiotensin receptor blockers (ARBs); time since symptom onset; percentage of lung affected on the CT scan; presence of confusion; presence of dehydration; respiratory frequency; oxygen saturation without oxygen; oxygen flow at inclusion; systolic blood pressure; lymphocyte count, neutrophil count, platelet count and CRP. All variables included in the propensity score model reflected knowledge available at baseline. Standardised differences were examined to assess balance, with a threshold of 10% designated to indicate clinically meaningful imbalance.^14^

The cumulative incidence of outcomes was computed by using the Kaplan-Meier method. Cox proportional hazards models were used to compute IPTW hazard ratios. IPTW estimates of the relative risk were computed for binary outcomes.

To account for immortal time bias, all patients from the no-CTC group who achieved the primary outcome (intubation or death) during the grace period were randomly assigned to one of the two groups, given that their observational data were compatible with both groups at the time of the event [22]. Assignment of these patients in the CTC group was based on their probability of receiving the intervention, accounting the moment patients met the primary outcome, by using a binomial distribution of parameters (n) the number of patients from the no-CTC group who met the primary outcome on day X (X going from 1 to 5) and (p) the probability of receiving corticosteroids after day X. Patients from the no CTC group lost to follow-up before the end of the grace period were assigned in the no-CTC group.

Outcomes are presented described in the total population and in the subgroups of patients with higher oxygen requirements at baseline (oxygen flow at baseline ≥3L/min), with severe inflammatory syndrome (CRP at baseline ≥ 100mg/L) and by time since symptom onset (≤ 7 days or > 7 days). In each subgroup, we recalculated the propensity score to balance the differences in baseline variables between treatment groups.

Because prescriptions of corticosteroids were based on clinicians’ decisions, not all patients eligible in the study would have been given corticosteroids. Therefore, we conducted a secondary analysis targeting the average treatment effect among the treated (ATT), which measured the effect of corticosteroids on patients who received them [18]. For that, we used a standardised mortality ratio weighting (SMRW), based on patients’ propensity score to balance groups at baseline[23].

For safety outcomes, all patients who received corticosteroids before Day 28, whatever the dose and timing, were considered in the CTC group.

To assess the robustness of findings, we evaluate how sensitive the results were to unmeasured confounding by the E-value, which measures the minimum strength of association an unmeasured confounder would need to have with both the treatment and the outcome to fully explain away the treatment effect [24]. We also performed sensitivity analyses. First, we conducted a trimmed analysis that was truncated at the region of common support, defined as the overlap between the range of propensity scores in the CTC group and the standard of care group. Patients with propensity scores outside the region of common support were excluded from this analysis. Second, to account for a potential centre effect, we computed the primary outcome by omitting one centre at a time. Third, we also assessed the impact of the 5 days grace period on our results by taking a shorter grace period of 48h. Fourth, we assessed another version of the intervention involving solely the addition of corticosteroids with at least 0.8 mg/kg/day eq. prednisone to standard of care. Patients who received a lesser dose, in addition to antivirals were dropped from analysis. Finally, since some patients received corticosteroids at a lower dose and/or subsequently received corticosteroids after 5 days, we specified an additional comparison mimicking an intention-to-treat analysis: all patients eligible for the study were analysed, and those whose data were not compatible with the CTC group were analysed in the control group.

Missing baseline and outcome variables were handled by multiple imputations by chained equations using the other variables available. All statistical analyses were performed with the R statistical package version 3.6.1 or later (The R Foundation for Statistical Computing, https://www.R-project.org/).

### Patient and public involvement

Neither patients nor the public were involved in the conception or conduct of the study.

## Results

### Patients and baseline characteristics

Among the 965 patients eligible for analysis, 194 received corticosteroids with at least 0.8 mg/kg/d eq. prednisone or 0,4 mg/kg/d eq. prednisone if co-administrated with lopinavir-ritonavir within five days from eligibility, 697 did not receive corticosteroids, 28 received corticosteroids at a dose less than 0.8 mg/kg/d eq. prednisone or 0,4 mg/kg/d eq. prednisone if co-administrated with ritonavir, and 46 received corticosteroids after five days from eligibility.

In our main analysis, to account for time-dependent bias, we randomly assigned patients from the control group who reached the primary outcome (intubation or death) during the grace period to one of the two groups, given that their observational data were compatible with both groups at the time of the event [22]; thus, we compared 203 participants in the CTC group to 688 in the standard of care group (**Figure 1**).

**Figure 1:**
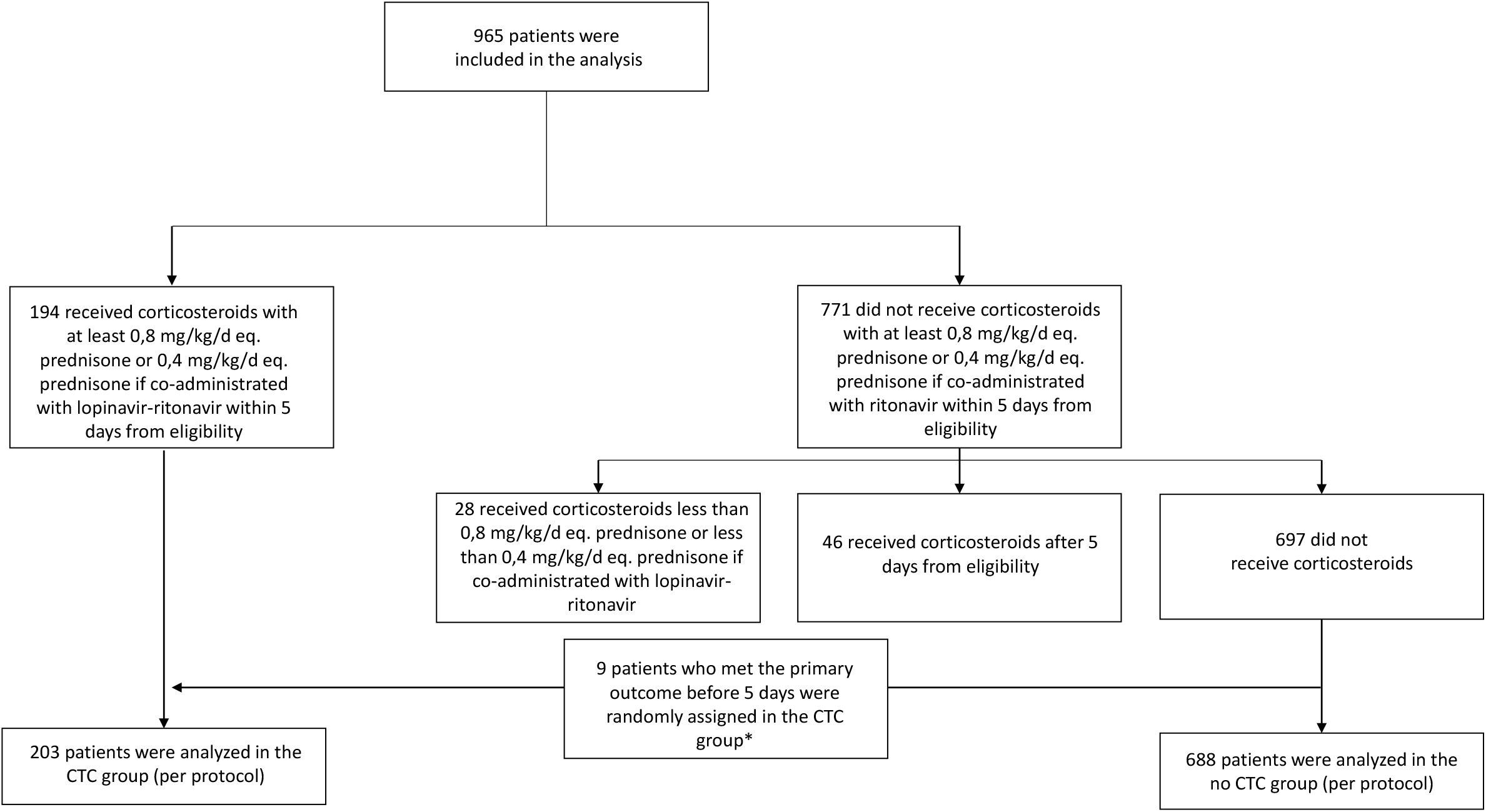
Study flow chart.

The patients’ median age was 63 years (interquartile range [IQR], 53 to 70 years), and 66.4% were men. The median time between symptom onset and eligibility was 8 days (IQR, 6 to 10 days). For 66% of patients, eligibility date was the date of hospitalisation. Overall, patients in the no-CTC group had less oxygen requirements with 22 (10.8%) and 130 (18.9%) patients receiving ≤ 1L/min oxygen at baseline. Among the 194 patients who received corticosteroids, 53 (27.3%) received dexamethasone, 71 (36.6%) methylprednisolone, 21 (10.8%) prednisolone and 49 (25.3%) prednisone. The median duration of treatment was 7 days, IQR 6 to 11. The median daily dose of corticosteroids (eq. prednisone) was 1.3 mg/kg/day, IQR 1.0 to 1.7. The median time between symptom onset and start of corticosteroids was 10 days, IQR 8 to 13. Among those patients, 81 (41.8%) received a high dose of corticosteroids (≥120 mg/day) [25]. Concerning other anti-COVID drugs prescribed to patients at baseline, 84 (9.4%) patients were treated with hydroxychloroquine and 60 (6.7%) were treated by lopinavir-ritonavir (**Table 1, Supplementary material 2**).

**Table 1:** Demographic and clinical characteristics of the patients in the CTC and no CTC group, at baseline. The number in brackets in the first column corresponds to the actual quantity of data available for the corresponding variables before imputation of missing baseline data by multiple imputations by chained equations using the other variables available. Abbreviations: IQR, interquartile range; NYHA, New York Heart Association; BMI, body mass index; ACEIs, angiotensin-converting enzyme inhibitors; ARBs, angiotensin receptor blockers. * 127 patients did not have a CT scan at admission; **Corresponds to the data of the 194 patients who received corticosteroids within 5 days of eligibility, with at least 0.8 mg/kg/day eq. prednisone or 0,4 mg/kg/day eq. prednisone if co-administrated with lopinavir-ritonavir. Results are presented as % (absolute number) unless stated otherwise.

| Characteristic | Total<br>(n=891) | CTC<br>group<br>(n=203) | No-CTC<br>group<br>(n=688) |
| --- | --- | --- | --- |
| **Demographic and clinical data** |  |  |  |
| Age, median (IQR) –year | 63 (53 – 70) | 64 (56 – 72) | 62 (52 – 70) |
| Male sex | 66.4 (592) | 71.4 (145) | 65.0 (447) |
| Comorbidities |  |  |  |
| Chronic respiratory disease (incl. asthma) | 5.6 (50) | 9.4 (19) | 4.5 (31) |
| Chronic heart failure (NYHA I-III) (n=890) | 3.5 (31) | 3.9 (8) | 3.3 (23) |
| Cardiovascular diseases (incl. hypertension) | 44.7 (398) | 48.3 (98) | 43.6 (300) |
| Diabetes not requiring insulin (n=886) | 17.6 (156) | 18.2 (37) | 17.4 (119) |
| Diabetes requiring insulin (n=889) | 5.8 (52) | 4.5 (9) | 6.3 (43) |
| Chronic kidney failure | 3.0 (27) | 3.4 (7) | 2.9 (20) |
| Liver cirrhosis | 0.4 (4) | 0.5 (1) | 0.4 (3) |
| Immunosuppression (n=890) | 5.4 (48) | 6.9 (14) | 4.9 (34) |
| BMI > 30 kg/m<sup>2</sup> (n=742) | 32.3 (240) | 27.8 (52) | 33.9 (188) |
| Treatment by ACEIs or ARBs (n=888) | 26.8 (238) | 30.7 (62) | 25.7 (176) |
| **COVID-19 data** |  |  |  |
| Time from symptom onset to eligibility, median (IQR) – days (n=887) | 8 (6 – 10) | 8 (6 – 11) | 8 (6 – 10) |
| Confusion at date of eligibility | 7.9 (70) | 10.3 (21) | 7.1(49) |
| Dehydration at date of eligibility (n=888) | 7.2 (64) | 8.9 (18) | 6.7 (46) |
| Respiratory rate, median (IQR) /min (n=739) | 24 (20 – 28) | 26 (21 – 30) | 24 (20 – 28) |
| Oxygen saturation (without oxygen), median (IQR) (n=795) | 93 (90 – 95) | 92 (88 – 95) | 93 (91 – 95) |
| Oxygen flow at admission, median (IQR) – L/min | 2.0 (2.0 – 4.0) | 3.0 (2.0 – 4.0) | 2.0 (2.0 – 3.0) |
| Systolic blood pressure, median (IQR) – mmHg (n=879) | 129<br>(116 – 140) | 129<br>(117 – 143) | 129<br>(115 – 140) |
| Neutrophils count, median (IQR) – /mm<sup>3</sup> (n=851) | 4790<br>(3435 – 6498) | 5000<br>(3508 – 6900) | 4750<br>(3400 – 6290) |
| Lymphocytes count, median (IQR) – /mm<sup>3</sup> (n=852) | 900<br>(690 – 1201) | 860<br>(642 – 1210) | 920<br>(700 – 1200) |
| Platelets count, median (IQR) – x1000/mm<sup>3</sup> (n=878) | 197<br>(157 – 254) | 182<br>(146 – 254) | 198<br>(159 – 253) |
| C-reactive protein (CRP) > 40 mg/l | 100 (64 – 149) | 111 (73 – 174) | 95 (61 – 141) |
| Percentage of lung affected > 50% on the CT scan (n=694*) | 17.1 (119) | 28.4 (50) | 13.3 (69) |
| **Treatment data** |  |  |  |
| Corticosteroid** |  |  |  |
| Dexamethasone | 53 (27.3) | 53 (27.3) | - |
| Methylprednisolone | 71 (36.6) | 71 (36.6) |  |
| Prednisolone | 21 (10.8) | 21 (10.8) |  |
| Prednisone | 49 (25.3) | 49 (25.3) |  |
| High dose corticosteroid ($\geq 120\text{mg/day}$) | 41.8 (81) | 41.8 (81) | - |
| Corticosteroid treatment duration, median (IQR) – days (n=194) | 7 (6 – 11) | 7 (6 – 11) | - |
| Time from symptom onset to treatment by corticosteroids, median (IQR) – days | 10 (8 – 13) | 10 (8 – 13) | - |
| Treatment by lopinavir-ritonavir at baseline (n=890) | 6.7 (60) | 15.3 (31) | 4.2 (29) |
| Treatment by hydroxychloroquine at baseline (n=889) | 9.4 (84) | 12.3 (25) | 8.6 (59) |

During follow-up, 146 patients received additional treatment with hydroxychloroquine; 111 received lopinavir-ritonavir; 14 received remdesivir; and 19 received interleukin-6 inhibitors (**Supplementary material 3**).

### Propensity score model development

Propensity scores ranged from 0.08 to 0.89 and from 0.04 to 0.83 in the CTC and no CTC groups, respectively, with 93.2% in the region of common support [0.08 – 0.83] (**Supplementary material 4**). Among the 25 covariates in the planned propensity score, one (liver cirrhosis) was dropped from the final model because only one patient had liver cirrhosis in the CTC group (compared with three in the no-CTC group). After applying IPTW, all 25 covariates (incl. liver cirrhosis) in the planned propensity score had weighted standardised differences below 10% (**Supplementary material 5**). After applying SMRW, 23 out of 25 covariates had weighted standardised differences below 10% (**Supplementary material 6**) while two, saturation without oxygen and chronic kidney disease had weighted standardised differences of 11%.

### Follow-up and outcomes

Among the 891 patients included in the main analysis, 78 had a follow-up less than 28 days (among whom 70 were discharged in good health status) and 63 died before Day 28. Median follow-up for patients alive was 80 days, interquartile range (IQR) 38 – 94.

In the unweighted sample, at Day 28, 18.0% (n=36) in the CTC group and 19.4% (n=131) in the no-CTC group had been intubated or died, hazard ratio 0.89, 95% confidence interval 0.62 to 1.28). Death occurred in 8.5% (n=17) of patients in the CTC group and 6.9% (n=46) in the no-CTC group, HR 1.23, 95% CI 0.70 to 2.13. At Day 28, 79.9% (n=148) of patients in the CTC group were weaned from oxygen, compared with 83.6% (n=522) in the no-CTC group (RR 0.96, 95% CI 0.88 to 1.03). Furthermore, 81.6% (n=151) patients in the CTC group were discharged to home/rehabilitation, compared with 84.4% (n=529) in the no-CTC group (RR 0.97, 95% CI 0.90 to 1.04).

In the IPTW analyses estimating the average treatment effect in the whole population, the cumulative rates of intubation or death were 20.1% in the CTC group, and 20.9% in the no-CTC group (wHR 0.92, 95% CI 0.61 to 1.39) (**Figure 2**). The cumulative death rates at Day 28 were 8.7% in the CTC group, versus 8.2% in the no-CTC group (wHR 1.03, 95% CI 0.54 to 1.98) (**Figure 3**). At Day 28, 80.8% of patients in the CTC group were weaned from oxygen, versus 82.5% in the no-CTC group (wRR 0.98, 95% CI 0.89 to 1.07). Further, 83.5% of patients in the CTC group were discharged to home/rehabilitation, versus 83.3 % in the no-CTC group (wRR 1.00, 95% CI 0.92 to 1.09) (**Table 2**).

**Table 2:** Primary and secondary outcomes at day 28. IPTW estimates the average treatment effect on the whole population (ATE). SMRW estimates the average treatment effect on the treated (ATT). CTC: corticosteroids-treated group. No CTC: standard of care, not treated with corticosteroids. IPTW: inverse probability of treatment weighting; ATE: average treatment effect; SMRW: standardised mortality ratio weighting; CI: confidence interval. *missing data were managed using multiple imputations by chained equations.

| Outcomes | No of events |  | Ratio (95% CI) | IPTW ratio* (95% CI) | SMRW Ratio* (95% CI) |
| --- | --- | --- | --- | --- | --- |
|  | CTC group (n=203) | No CTC group (n=688) |  |  |  |
| Intubation or death | 36 | 131 | HR 0.89<br>(0.62 – 1.28) | wHR 0.92<br>(0.61 – 1.39) | wHR 0.64<br>(0.43 – 0.97) |
| Death | 17 | 46 | HR 1.23<br>(0.70 – 2.13) | wHR 1.03<br>(0.54 – 1.98) | wHR 0.65<br>(0.33 – 1.30) |
| Oxygen weaning | 148/186* | 522/625* | RR 0.96<br>(0.88 – 1.03) | wRR 0.98<br>(0.89 – 1.07) | wRR 1.02<br>(0.92 – 1.13) |
| Discharge from hospital to home or rehabilitation | 151/186* | 529/627* | RR 0.97<br>(0.90 – 1.04) | wRR 1.00<br>(0.92 – 1.09) | wRR 1.02<br>(0.93 – 1.13) |

**Figure 2:**
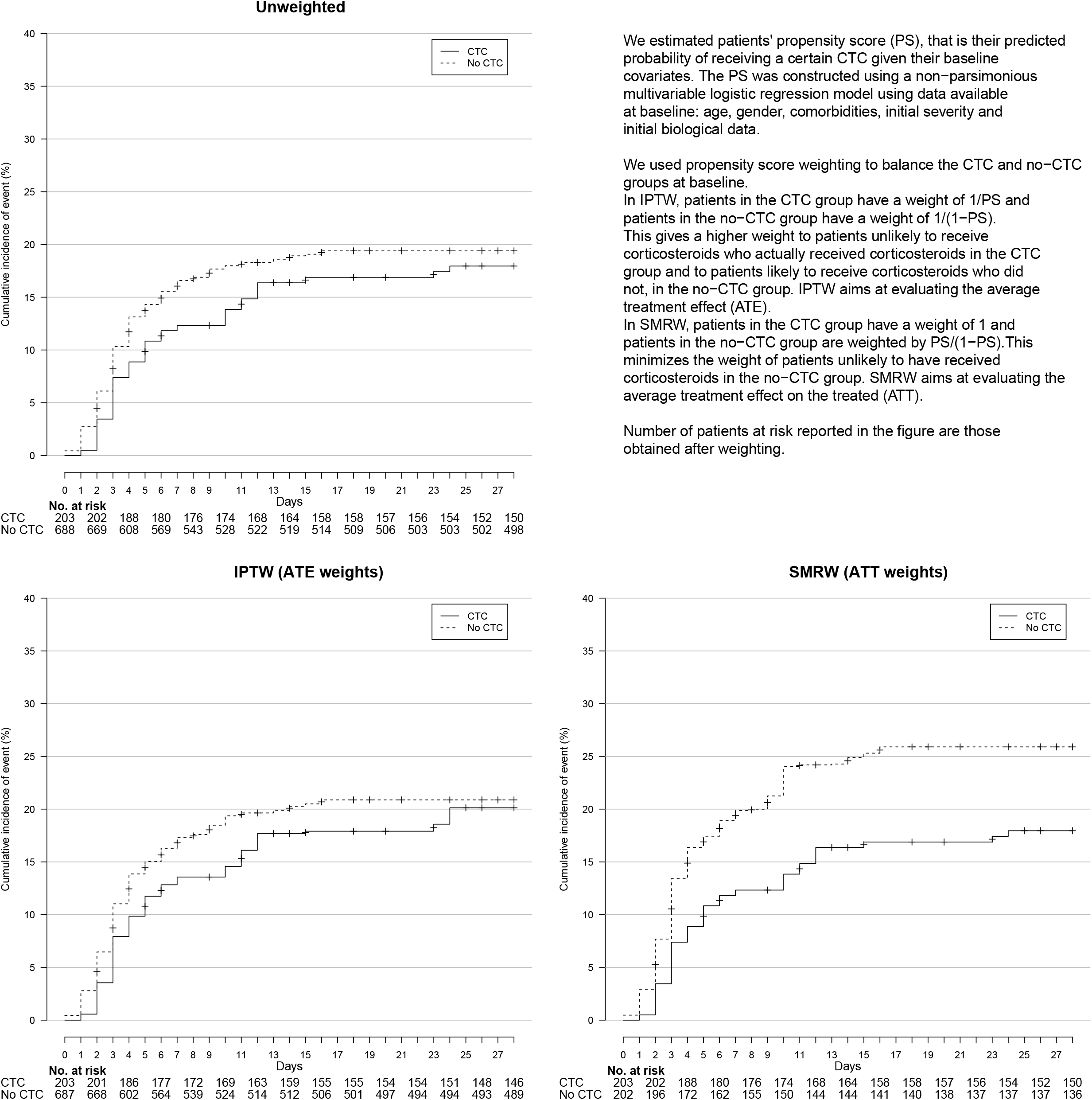
**Cumulative event curve for intubation or death** in the unweighted sample (top panel) IPTW sample (bottom-left panel) and SMRW sample (bottom-right panel).

**Figure 3:**
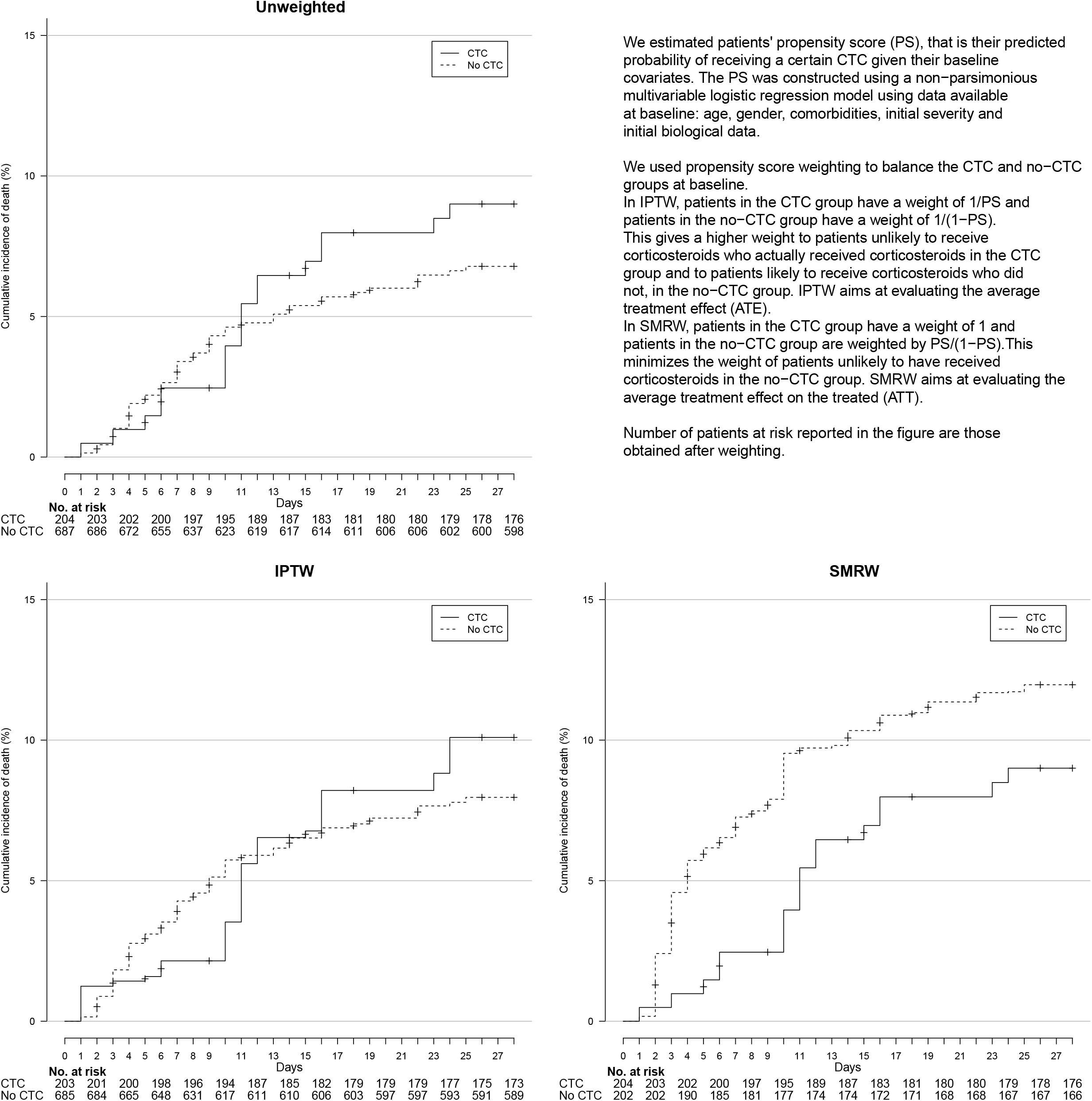
**Cumulative event curve for death** in the unweighted sample (top panel) IPTW sample (bottom-left panel) and SMRW sample (bottom-right panel).

In the SMRW analyses estimating the average treatment effect on the treated, the cumulative rates of intubation or death were 18.0% in the CTC group versus 25.9% in the no-CTC group (wHR 0.64, 95% CI 0.43 to 0.97, E-value: 2.05) (**Figure 2**). Cumulative death rates were 8.5% in the CTC group versus 12.5% in the no-CTC group (wHR 0.65, 95% CI 0.33 to 1.30) (**Figure 3**). At day 28, 79.9% of patients in the CTC group were weaned from oxygen, versus 78.5% in the no-CTC group (wRR 1.02, 95% CI 0.92 to 1.13). Further, 81.6% of patients in the CTC group had been discharged to home/rehabilitation, versus 79.6% in the no-CTC group (wRR 1.02, 95% CI 0.93 to 1.13) (**Table 2**).

Results in subgroups are presented in **Figure 4, Supplementary material 7 and 8**. In both IPTW and SMRW analyses, for patients with a baseline oxygen flow ≥ 3 L/min, corticosteroids were associated with a significant reduction of the incidence of the primary outcome for patients with a baseline oxygen flow ≥ 3 L/min (wHR 0.50, 95% CI 0.30 to 0.85 [E-value 2.59] and wHR 0.56, 95% CI 0.33 to 0.95 [E-value 2.35] for IPTW and SMRW analyses respectively). In both analyses, for patients with CRP>100mg/L, corticosteroids were associated with a significant reduction of the incidence of the primary outcome (Whr 0.44, 95% CI 0.23 to 0.85 [E-value 2.93] and wHR 0.41, 95% CI 0.22 to 0.75 [E-value 3.10] for IPTW and SMRW analyses respectively). For these patients, we also found a significant reduction in death rate (wHR 0.25, 95% CI 0.10 to 0.65 [E-value 4.51] and wHR 0.30, 95% CI 0.11 to 0.81 [E-value 4.01] for IPTW and SMRW analyses respectively).

**Figure 4:**
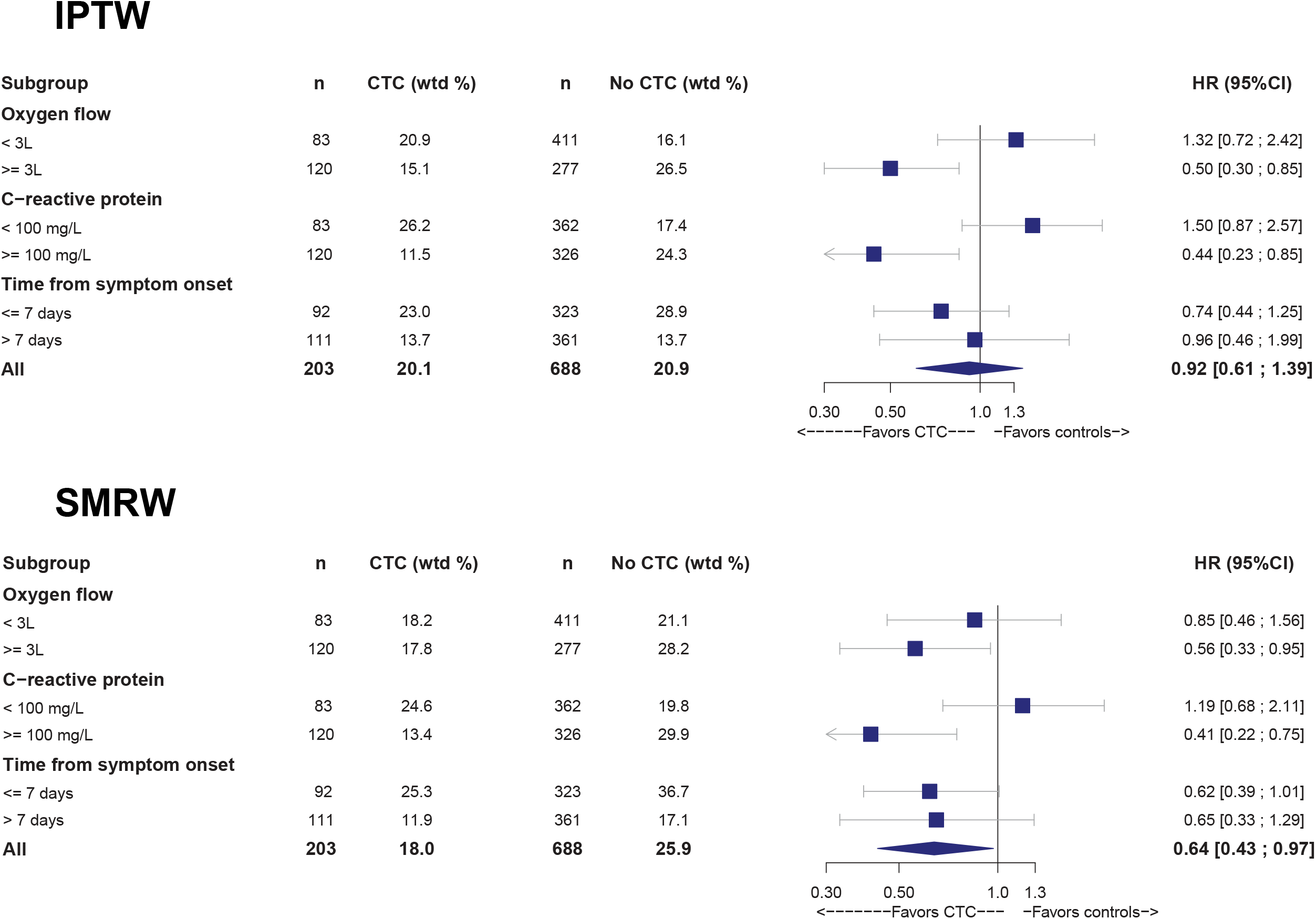
Effect of corticosteroids on 28-Day intubation or death in subgroups defined by oxygen requirement, levels of C-reactive protein and time since symptom onset, at baseline.

Sensitivity analyses on the trimmed sample were consistent with the principal analysis for both IPTW and SMRW analyses (**Supplementary material 9**). We found no significant centre effect in our data (S**upplementary material 10**). Results were similar in a secondary analysis with a shorter grace period of 48hours (wHR 0.84, 95% CI 0.49 to 1.43 and wHR 0.57, 95% CI 0.35 to 0.95 for the primary outcome, in IPTW and SMRW respectively) (**Supplementary data 11**). Focusing on the efficacy and safety of corticosteroids at 0.8 mg/kg/day eq. prednisone and dropping patients who received a lesser dose in addition to lopinavir/ritonavir from analysis did not change results (wHR 1.03, 95% IC 0.66 to 1.61 and wHR 0.66, 95% IC 0.43 to 1.00 for the primary outcome, in IPTW and SMRW respectively) (**Supplementary data 12**). When adding the patients who received corticosteroids at a dose lower than 0.8mg/kg/day (or 0,4 mg/kg/day eq. prednisone if co-administrated with lopinavir-ritonavir) and those who received corticosteroids after five days from eligibility in the no CTC group (mimicking an intention-to-treat analysis), we did not retrieve the association between treatment by corticosteroids and the primary outcome in SMRW analyses (wHR 1.00, 95% CI 0.67 to 1.48 and wHR 0.78, 95% CI 0.53 to1.15 for the primary outcome, in IPTW and SMRW respectively) (**Supplementary data 13**).

### Safety

Overall, 464 adverse events were extracted from electronic health records (150/283 (53.0%) among patients who received corticosteroids and 314/682 (46.0%) among patients who did not) (**Table 3**). Difference involved mainly a higher number of hyperglycaemia events in patients treated by corticosteroids (64/283 [22.6%] vs. 86/682 [12.6%]). There was no increased rate of infection in patients who received corticosteroids (50/283 [17.7%] among patients who received corticosteroids vs. 128/682 [18.8%] among patients who did not). Similar results were found for ventilator-associated pneumonia (17/283 [6.0%] for patients who received corticosteroids vs. 61/682 [8.9%]).

**Table 3:** Adverse events. Adverse events are counted in the safety population, without weighting. Results are presented as % (absolute number).

| Adverse event | CTC group<br>(n=283) | No-CTC group<br>(n=682) | Difference<br>(95%CI) |
| --- | --- | --- | --- |
| Any | 53.0 (150) | 46.0 (314) | 7.0 (0.0 ; 13.9) |
| **Expected with corticosteroids** |  |  |  |
| Infection (incl. Ventilator-associated pneumonia) | 17.7 (50) | 18.8 (128) | -1.1 (-6.4 ; 4.2) |
| Ventilator-associated pneumonia | 6.0 (17) | 8.9 (61) | -2.9 (-6.4 ; 0.6) |
| Hyperglycaemia | 22.6 (64) | 12.6 (86) | 10.0 (4.5 ; 15.5) |
| Hypertension | 10.6 (30) | 11.1 (76) | -0.5 (-4.8 ; 3.8) |
| Confusion or psychiatric manifestation | 1.4 (4) | 1.9 (13) | -0.5 (-2.2 ; 1.2) |
| Atrial fibrillation | 4.6 (13) | 3.8 (26) | 0.8 (-2.0 ; 3.6) |
| Hypokalaemia or fluid overload | 1.1 (3) | 1.3 (9) | -0.3 (-1.7 ; 1.2) |
| **Other severe adverse events** |  |  |  |
| Thromboembolic event (incl. pulmonary embolism) | 2.8 (8) | 3.5 (24) | -0.7 (-3.1 ; 1.7) |
| Pulmonary embolism | 2.1 (6) | 2.6 (18) | -0.5 (-2.6 ; 1.5) |
| Increased serum levels of aspartate aminotransferase | 5.3 (15) | 4.4 (30) | 0.9 (-2.1 ; 3.9) |
| Renal failure | 2.5 (7) | 2.6 (18) | -0.2 (-2.3 ; 2.0) |

## Discussion

We report a multicentre observational study from 51 general and university hospitals that used real-world data collected from routine care to assess the efficacy and safety of corticosteroids in 965 consecutive patients hospitalised for COVID-19 hypoxemic pneumonia with systemic inflammation. In our main analysis that estimated the average treatment effect, we did not find a significant difference in the rate of intubation or death nor death between patients who received corticosteroids at ≥ 0.8 mg/kg/d eq. prednisone (or 0,4 mg/kg/d eq. prednisone if co-administrated with lopinavir-ritonavir) and those who did not although the result was compatible with a 40% reduction in hazard. Results were unchanged if we focused solely on patients who received corticosteroids at ≥ 0.8 mg/kg/d eq. prednisone. The rate of patients discharged from hospital or weaned from oxygen did not decrease either. In a secondary analysis targeting the average treatment effect on the treated (ATT), which minimises the weight of patients unlikely to have received corticosteroids in the no-CTC group, adding corticosteroids to the standard of care was associated with a significant reduction in the incidence of intubation or death at 28 days. These results are consistent with the average treatment effect (ATE) observed in the subgroups analyses targeting patients with higher oxygen needs or patients with a CRP>100mg/L at baseline. The difference between these two analyses suggest that physicians chose to prescribe corticosteroids to a population more severe than those strictly defined by the inclusion criteria of the study. Within the population treated by physicians, our results are consistent with those from the RECOVERY trial and support the use of corticosteroids to reduce intubation or death. For patients less severe, our results question the benefit risk balance of corticosteroids. Especially, although the safety, data from our study were reassuring regarding secondary bacterial or fungal infections, we noted only a higher proportion of hyperglycaemia occurrences in patients treated by corticosteroids.

Differences between results from the RECOVERY trial and those from our study may be explained by several factors. First, our study was based on exhaustive data from consecutive patients hospitalised for COVID-19 pneumonia and who met the inclusion criteria. Thus, our analyses involved a large number of less severe patients who had limited requirements in oxygen (mainly in the no-CTC group with 18% of patients who received ≤1L/min oxygen at baseline) and whose condition may be closer to the subgroup of patients from the RECOVERY trial who were not under oxygen at the time of randomization. Second, our population differs because we included only patients ≤ 80 years old hospitalised in conventional wards and excluded patients with severe chronic conditions such as deep undernutrition; renal diseases requiring dialysis; chronic heart failure NYHA IV; liver cirrhosis Child C and chronic respiratory insufficiency under oxygen therapy before admission. In contrast, >20% of patients from the RECOVERY trial were aged 80 or older. This may explain the difference in death rates at 28 days in the no-CTC groups (8.2% in our study and 25% in the RECOVERY trial). Third, the median time from illness onset to corticosteroid therapy is also a key issue. In our main analysis, the median time from onset of illness to initiation of corticosteroids was 10 days, IQR 8 to 13, and later than in the RECOVERY trial (8 days, IQR 5 to 13) [12]. Fourth, in our study, the main corticosteroids used were dexamethasone and methylprednisolone. Methylprednisolone has a lesser mineralocorticoid activity while dexamethasone possesses a higher glucocorticoid activity. Theoretically, methylprednisolone has the advantage of parenteral administration, a quicker onset of action and a shorter duration of action compared to the dexamethasone [26]. In addition, risk of long-term side effects like fluid retention, hypokalemia, hypercortisolism and hyperglycemia are less likely with methylprednisolone.

Besides corticosteroids, other immunomodulatory drugs strategies that block hyper-inflammation are under evaluation. Preliminary results of interleukin-6 or interleukin-1 blockade and/or anti-TNF are in favour of a beneficial effect of immunomodulatory drugs during the inflammatory phase of COVID-19 infection [27,28], and the confirmatory results of the CORIMUNO-TOCI and RECOVERY randomised controlled trials are upcoming. However, in a large outbreak context, corticosteroids are largely more easily available than other immunomodulators such as interleukin receptor inhibitors. They are commonly used by a large spectrum of clinicians and can be easily considered and deployed in low- and middle-income countries, or in elderly health care facilities.

### Strengths and Limitations of study

One of the major strengths of this study is the analysis of a large number of consecutive patients, from a broad variety of centres in France and Luxembourg. Another strength is that all safety data have been reviewed in double and independently by several clinicians which did not note any warning signal for using corticosteroids in COVID-19 pneumonia in terms of secondary bacterial or fungal infection, even for ventilator-associated pneumonia. In terms of adverse events, there were only a higher number of hyperglycaemia occurrences in patients treated by corticosteroids which are usually transient and reversible with the stoppage of the steroids.

However, our study has several limitations. First, despite the use of robust methods and statistical techniques to draw causal inferences. our study is observational, and potential unmeasured confounders may bias our results. Second, our study used real world data, with a heterogeneity in the prescription of corticosteroids treatment in terms of drugs, time of start, dose and duration. Third, we were not able to analyse the impact of the duration of corticoid prescription as we dealt with observational data collected from routine care. For example, some patients only received three days of corticosteroids because an event occurred on the fourth day. Only trials where the dose and duration are specified before treatment, in “intent-to-treat”, can answer this question. Fourth, identification of patients eligible in the study was performed manually with a patient-by-patient screening of all patients hospitalized in the study centres within the study dates. As we used data from a large number of hospitals using different electronic record system, we were not able to standardize this process and this may have led to missing some patients. Fifth, we did not adjust for different methods to collect biological data such as C-reactive protein. This choice was based on the pragmatic consideration that values of CRP from one hospital are usually considered valid by physicians from another hospital for taking decisions on patients’ care, however, this may have generated imprecision in our analyses. Sixth, our sample was limited to the number of eligible patients available at the time of analysis; we cannot rule out the possibility that our findings are due to a lack of power, especially regarding the subgroups analysed, with multiple findings showing a trend towards benefit. Seventh, our subgroup analysis by time since symptom onset is only a proxy of patients’ immune status. Further analyses should investigate the right timing of corticosteroids as a function of the immune phenotypes of patients. Finally, since we included only patients < 80 years old, we cannot reach a conclusion about the possible efficacy of steroids in preventing severe forms of the disease in elderly.

## Conclusions

We found no association between the use of corticosteroids and intubation or death for all patients <80 years old hospitalised for COVID-19 pneumonia in non-ICU settings. However, use of corticosteroids was associated with a reduction in rate of intubation or death for patients actually treated in real life, those with higher oxygen requirements (≥ 3L/min) and those with a severe inflammatory syndrome (CRP ≥ 100mg/L) at baseline. These data support the support the use of corticosteroids for patients with mild COVID; future studies need to confirm the right timing and to determine the best dose and duration of this treatment

## Data Availability

The data are available on request.

## Ethical approval

This study conformed to the ethical guidelines of the 1975 Declaration of Helsinki and was approved by the ethics committee of the Henri-Mondor Hospital (AP-HP), France (number 00011558) and by the National ethics committee of Luxembourg (number: 0620-101).

## Data sharing

The data are available on request.

## Dissemination to related patient and public communities

We plan to issue a press release on official publication of this manuscript and disseminate our findings through social media outlets, to ensure the results of the study have a broad public outreach.

The lead author (X. Lescure) affirms that the manuscript is an honest, accurate, and transparent account of the study being reported; that no important aspects of the study have been omitted; and that any discrepancies from the study as planned have been explained.

## Contributorship statement

V-TT, EP, PR and F-XL conceived the study; MM, FB, OB, TP, LG, FG and F-XL participated in data collection; V-TT, EP and PR performed the statistical analyses; MM, OB, TP, FG and F-XL reviewed the data for the adverse events; MM, LG, and F-XL provided administrative and logistic supports; V-TT, MM and F-XL wrote the first draft of the manuscript; All members of the writing committee contributed to the writing of the manuscript. All members of the writing committee meet the ICMJE criteria. All members of the writing committee agree with the manuscript results and conclusions. F-XL is the guarantor, had full access to the data in the study, and takes responsibility for the integrity of the data and the accuracy of the data analysis.

All collaborators are listed in the supplementary appendix.

